# Toward multimodal MRI biomarkers of PTSD: functional and structural connectivity signatures in WTC responders

**DOI:** 10.64898/2026.07.13.26357932

**Authors:** Azzurra Invernizzi, Davide Folloni, Elza Rechtman, Alison C. Pellecchia, Stephanie Santiago-Michels, Evelyn J. Bromet, Roberto G. Lucchini, Benjamin J. Luft, Sean A. Clouston, Cheuk Y. Tang, Megan K. Horton

## Abstract

**Background:** Post-traumatic stress disorder (PTSD) remains highly prevalent affecting ~23% of World Trade Center (WTC) responders more than two decades after 9/11. While MRI studies have identified neural differences associated with PTSD, these findings have not translated into improved treatment. We introduce a novel multimodal MRI approach, DAta-driven Network Connectivity Estimate (DANCE), integrating structural and functional magnetic resonance imaging (MRI) to better capture PTSD mechanisms and inform biomarkers.

**Methods:** In 96 WTC responders, including 45 with current WTC-related PTSD and 51 without PTSD. We applied graph theory to resting-state functional MRI to identify functional hubs via eigenvector centrality and identified divergence between groups using partial least squares discriminant analysis (PLS-DA). From diffusion MRI, we reconstructed five anatomical tracts (i.e., streamlines) in the temporal lobes. Using DANCE, we quantified the differential distribution of streamlines of the reconstructed tracts connecting the functional hubs. We then tested whether WTC exposure duration moderated associations between PTSD and DANCE indices.

**Results:** Responders with PTSD showed altered centrality in nine functional hubs (AUC=0.75 (0.651-0.847)) including bilateral anterior inferior temporal gyrus, right superior parietal lobule, right anterior parahippocampal gyrus, right anterior/posterior superior temporal gyrus (STG), right caudate nucleus, left amygdala and brainstem. Connectivity differences emerged in four tracts: hippocampus, parahippocampus, inferior and superior temporal gyri (STG). DANCE differed in the inferior fronto-occipital fasciculus (IFOF), medial (IFLmed) and lateral (IFLlat) components of the inferior longitudinal fasciculus and in the middle longitudinal fascicle (MdLF). WTC exposure duration significantly moderated the association between PTSD and DANCE values in the IFLmed, right posterior STG (p= 0.035).

**Conclusion:** Our novel DANCE approach revealed converging functional and anatomical connectivity alterations uniquely associated with PTSD in WTC responders and offers compelling evidence for distinct neurobiological signatures of the disorder. These findings significantly advance our understanding of PTSD pathophysiology and highlight potential biomarkers for diagnosis and targeted intervention.

## Introduction

Responders involved in the rescue and recovery efforts following the 9/11 attacks on the World Trade Center (WTC) were exposed to a complex mixture of toxic agents, arduous working conditions, and traumatic psychosocial stressors including fear for personal safety, injury or illness and, exposure to human remains^1^. Over twenty years after the WTC tragedy, ~23% of WTC-responders still experience clinically significant post-traumatic stress disorder (PTSD)^2–4^. PTSD is a psychiatric condition characterized by persistent and intrusive memories, nightmares, flashbacks, heightened emotional arousal, avoidance of stimuli or experiences associated with stressful events. While psychotherapeutic and pharmacological PTSD treatments are available, many patients do not improve following them, prematurely stopping or never initiating treatment^5^. This highlights the urgent need to understand the neural circuitry underlying this disorder to guide treatment and develop more individualized mechanism-based treatment options.

Recent neuroimaging studies of WTC-responders, including studies in our group, leverage structural or functional MRI to investigate brain changes and functional neural-mechanisms underlying this disorder^9–11^. Anatomical differences between WTC-responders with PTSD (WTC-PTSD) and those without PTSD (non-PTSD) include reduced cortical complexity across brain areas (frontal, parietal, and temporal)^10^. These structural changes were associated with increased PTSD symptom severity (re-experiencing, avoidance, hyperarousal, negative thoughts). Using resting-state fMRI (rs-fMRI), we previously identified significant differences in functional neuro-profiles of WTC-responders with and without PTSD in nine brain areas defined as functional hubs^11^.

Among the responders with PTSD, the duration of WTC exposure (i.e., months at the rescue and recovery site) was associated with increased changes in local functional connectivity within the right anterior parahippocampal gyrus and the left amygdala. Both regions are associated with PTSD^12–16^. The anatomical and functional findings in this WTC cohort confirmed previous literature suggesting that PTSD follows the ‘fear-conditioning’ paradigm ^17–19^ characterized by exaggerated amygdala responses and reduced functionality in frontal lobe and hippocampal regions^19–22^. While informative, these previous studies have relied largely on either anatomical or functional MRI metrics, sometimes providing discrepant findings and limiting the identification of robust biomarkers of PTSD. This raises a central question: do observed alterations reflect structural differences, functional dysregulation, or both? And if both, which neural hubs are most consistently affected?

Here, we implement a machine learning multimodal approach leveraging anatomical and functional MRI metrics that offers a novel and accurate assessment of neural mechanisms underlying PTSD. By combining information about the white matter anatomical structures and gray matter functional data, we aim to overcome some of the limitations inherent of each MRI modality used (i.e., limited anatomical contrast in rs-fMRI data)^6–8^ and estimate subject-specific connectivity fingerprints to provide a better understanding of brain impairment while integrating the important information from these MRI modalities. Our overarching goal is to identify neural connectivity biomarkers that reflect integrated structured and functional alterations associated with the traumatic events of 9/11. In this study, we integrated graph-based network metrics derived from rs-fMRI data with anatomical connectivity features derived from DTI data, leveraging data-driven statistical methods to generate a multimodal connectivity profile defined using DAta-driven Network Connectivity Estimate (DANCE) ^23–25^. We further explored the association between WTC exposure duration and DANCE.

## Results

### Demographic and clinical characteristics

Clinical and demographic characteristics of WTC responders with and without PTSD are reported in Table 1. Responders were on average 55.8 ± 5.3 years old and predominantly male (79%). By design, groups were matched on age at the time of scan, sex, race/ethnicity, and education. Major Depression Disorder (MDD), daily psychotropic medication use, and PTSD symptoms significantly differ between groups.

**Table 1.** Sociodemographic and clinical characteristics of WTC responders in the current study (N=96).

| Characteristics | WTC responders (n=96) | PTSD- (n=51) | PTSD+ (n=45) | p |
| --- | --- | --- | --- | --- |
| **Age (years)** |  |  |  | 0.383 |
| mean ± sd | 55.81 ± 5.26 | 56.25 ± 5.34 | 55.31 ± 5.18 | |
| **Sex (n, %)** |  |  |  | 0.827 |
| Male | 76 (79%) | 39 (51%) | 37 (49%) |  |
| Female | 20 (21%) | 12 (60%) | 8 (40%) |  |
| **Comorbidities (n, %)** |  |  |  |  |
| Major Depressive Disorder |  |  |  | <0.001* |
| Yes | 18 (19%) | 0 (0%) | 18 (40%) |  |
| **Medications (n, %)** |  |  |  |  |
| Psychotropic |  |  |  | <0.001* |
| No | 74 (77%) | 47 (92%) | 27 (60%) |  |
| Yes | 22 (23%) | 4 (8%) | 18 (40%) |  |
| Opioid |  |  |  | 0.643 |
| No | 92 (96%) | 49 (96%) | 43 (95%) |  |
| Yes | 4 (4%) | 2 (4%) | 2 (5%) |  |
| **WTC Exposure (mean ± sd)** |  |  |  | 0.853 |
| Total months on site | 2.63 ± 5.1 | 2.54 ± 5.2 | 2.73 ± 4.96 | |

Mean, standard deviation (sd), range (minimum and maximum values), and percentage (%) are reported. P-values quantify differences between WTC-PTSD and non-PTSD responders and the noted characteristics; they were derived using Student t-tests for continuous variables and $\chi^2$ tests for categorical variables. \*denotes significance of $p < 0.005$ .

### Functional neuroprofile in WTC responders

PLS-DA identified distinct functional neuro-profiles between WTC responders with and without PTSD, with the first discriminant component accounting for 18% of the variance in functional connectivity data (Figure 1). The ROC curve for the first discriminant axis demonstrated moderate classification performance (AUC = 0.749, 95% CI:0.651-0.847; Figure 2B). Nine brain areas contributed most to differentiating WTC-PTSD from non-PTSD responders, including the bilateral anterior inferior temporal gyrus, right superior parietal lobule, right anterior parahippocampal gyrus, right anterior and posterior temporal gyrus cortex, right caudate nucleus, left amygdala, and brainstem. These regions, hereafter referred to as “hubs,” were predominantly right-lateralized. The same hubs were identified by Invernizzi et al. using a standard permutation test, supporting their robustness (Figure 3).

**Figure 1.**
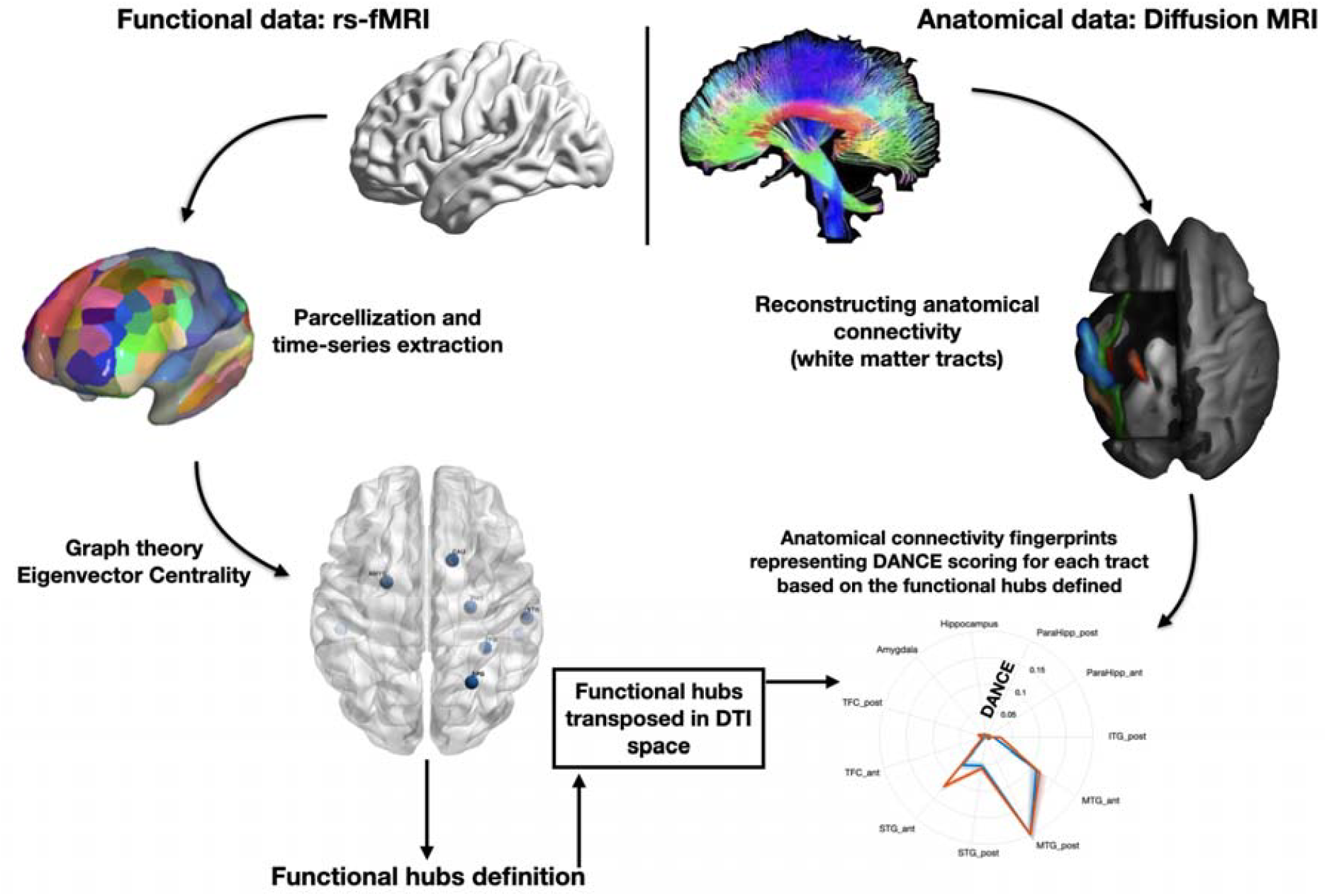
Functional and anatomical MRI data processing flowchart to generate DANCE, a multi-modal approach. Functional data: Resting-state fMRI data were preprocessed and the averaged time-series extracted for each Region-of-Interest (ROI) using the Harvard-Oxford atlas (111 cortical and subcortical brain areas). Eigenvector centrality (EC) was computed for each brain area. Using the EC values from 111 ROIs as inputs, PLS-DA was performed to model differences in local functional connectivity between WTC-PTSD and non-PTSD responders and identify functional hubs. **Anatomical data:** based on the identified functional hubs, five white matter tracts were reconstructed for each subject: the temporal branch of the cingulum bundle (CB), the medial (ILFmed) and lateral (ILFlat) projections of the inferior longitudinal fascicle, the middle longitudinal fascicle (MdLF) and the inferior-fronto-occipital fascicle (IFOF). The **DAta-driven Network Connectivity Estimate (DANCE)** was created based on the differential distribution of streamlines of the reconstructed tracts connecting the functional hubs and compared between WTC-PTSD and non-PTSD.

**Figure 2.**
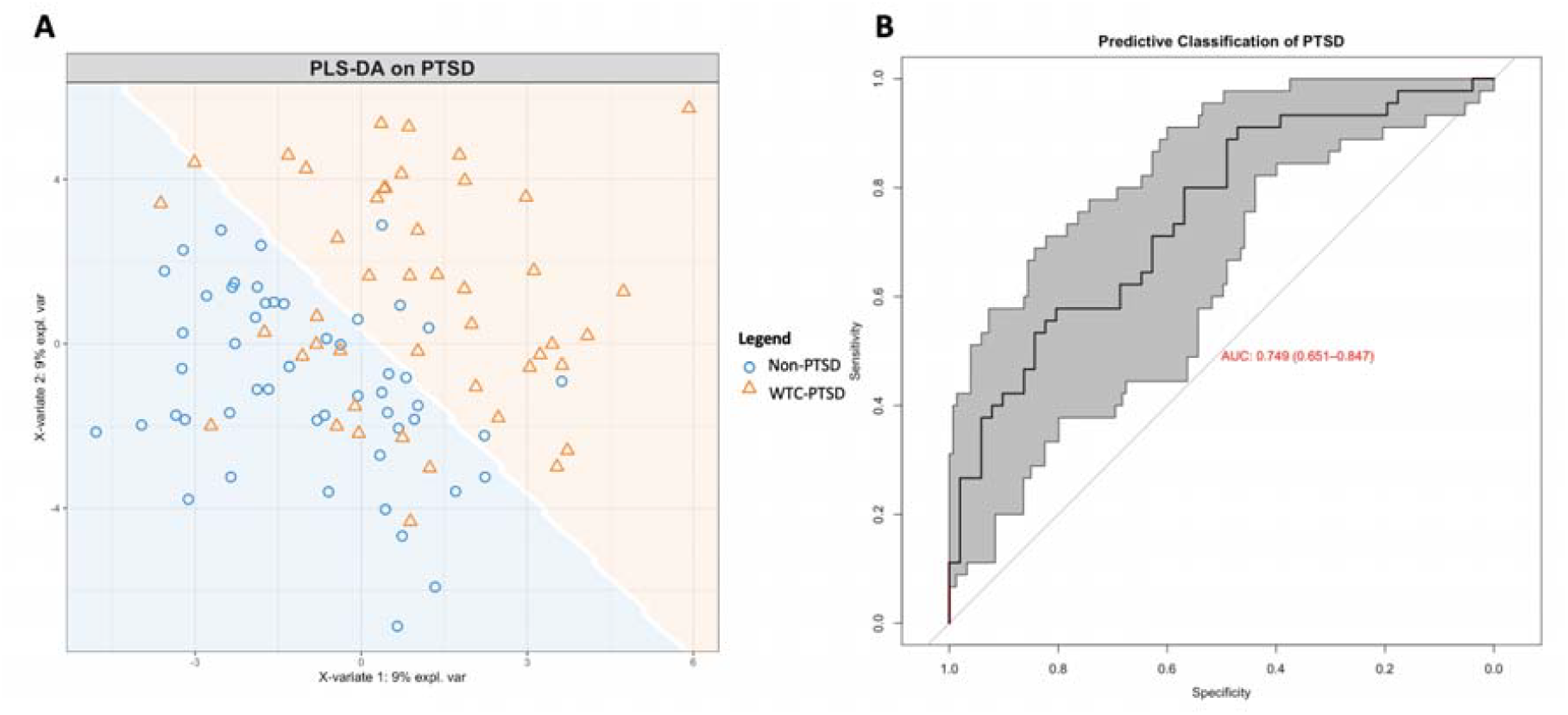
Partial least squares discriminant analysis (PLS-DA) shows clear and distinct functional connectivity patterns in WTC-PTSD and non-PTSD. Using the eigenvector centrality (EC) values from 111 brain regions as inputs, the PLS-DA constructs a lower-dimensional subspace that maximizes the separation between WTC-responders with and without PTSD. Panel A: Orange triangle (WTC-PTSD) and blue dots (Non-PTSD) show individual participants scores on the first (x-axis) and second (y-axis) discriminate axes derived through PLS-DA. Separation of orange triangle and blue dots (white line) illustrates the prediction area associated with each class, with subjects scoring in orange space predicted to be WTC-PTSD, those in blue space are predicted to be Non-PTSD. Panel B: Receiver operating characteristic (ROC) curve illustrating the sensitivity and specificity for predicting WTC-PTSD from the first discriminant axis with varying classification thresholds. To quantify uncertainty in the ROC curve and discriminant prediction, a confidence interval of the ROC curve was computed using 2000 stratified bootstraps, yielding an area-under-the-curve (AUC) of 0.749 (0.651-0.847).

**Figure 3.**
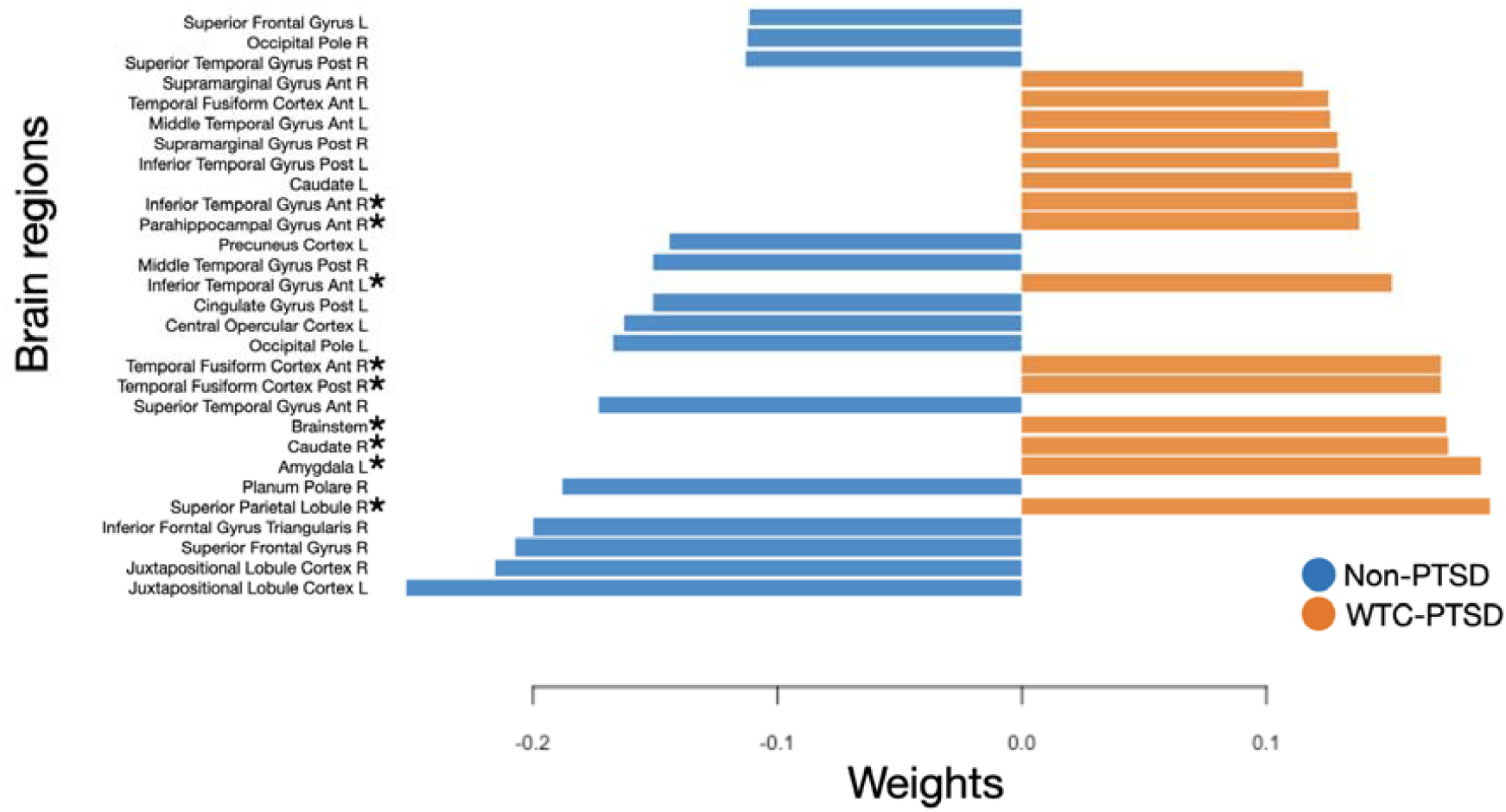
Partial least squares discriminant analysis (PLS-DA) loadings contributing to the divergence of WTC-PTSD from non-PTSD. For illustrative purposes, the figure shows the PLS-DA loading of the top 25% brain areas on a given dimensional subscale to determine which regions contribute most to the overall divergence between WTC-PTSD and non-PTSD responders. Loadings are color coded to indicate the group with a higher mean score on that variable, blue lines indicate variable loadings non-PTSD, and orange lines indicate variable loadings in WTC-PTSD. The loading direction (positive versus negative) indicates the direction of the associations between individual brain regions and PTSD. *Indicates p < 0.05 in brain areas where EC values differ significantly between groups. Complete loadings can be found in supplementary material (Figure S1).

### Anatomical connectivity neuroprofile in WTC responders

To quantify white matter connectivity to the altered functional hubs (DANCE profile), we derived tract-specific connectivity fingerprints (Figure 4 and S2)^23,46^. Five most relevant tracts were examined^11^: cingulum bundle (CB), medial (ILFmed) and lateral (ILFlat) projections of the inferior longitudinal fascicle, middle longitudinal fascicle (MdLF) and inferior-fronto-occipital fascicle (IFOF). Upon visual inspection, fingerprints shape and intensity differ between left–right hemisphere in CB and IFOF fingerprints (Figure 4C, 4E and Supplementary Figure S2). Hemispheres were analyzed separately.

**Figure 4.**
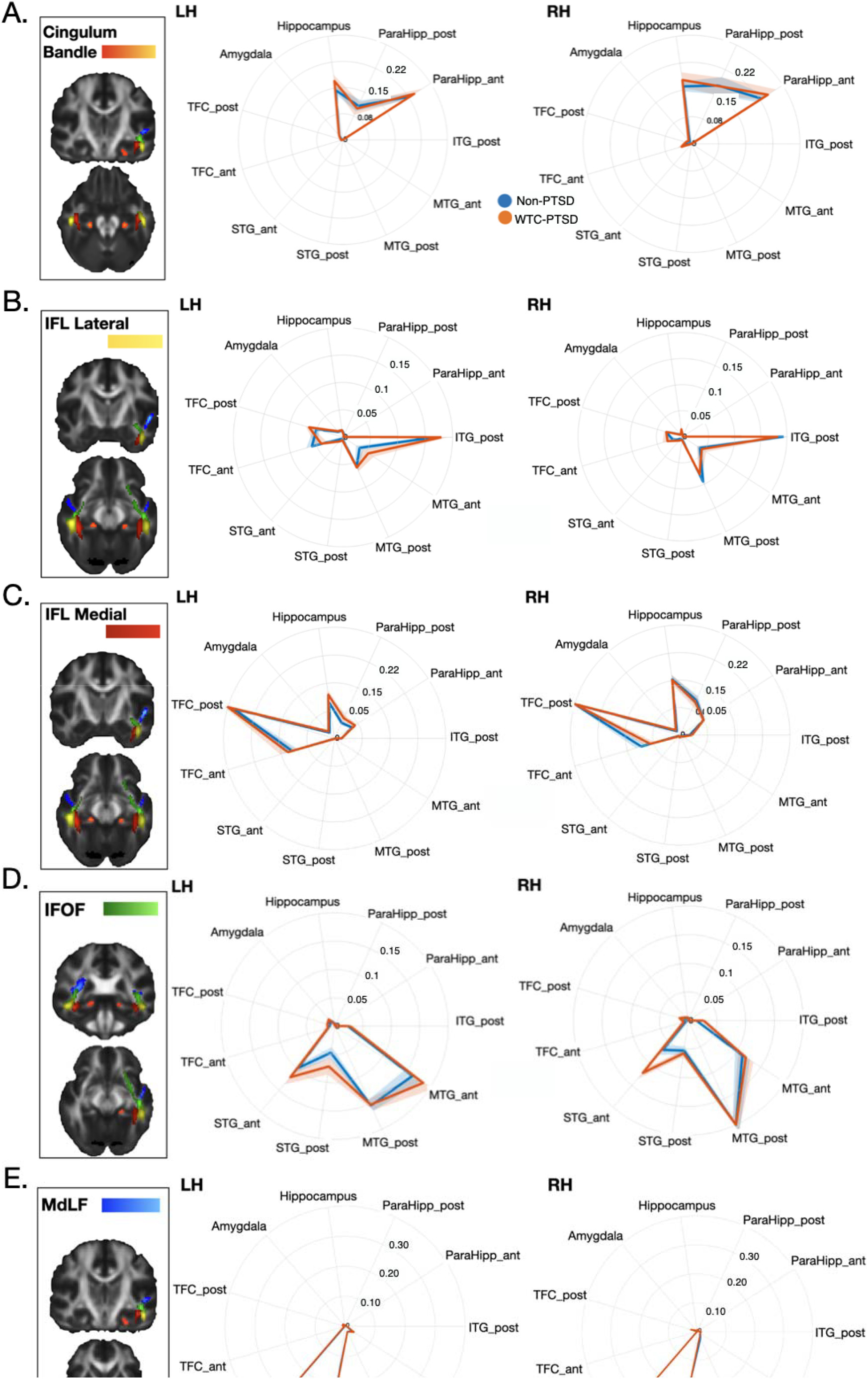
Comparison of anatomical connectivity fingerprints of the CB, ILFlat, ILFmed, IFOF and MdLF white matter tracts in WTC responders with and without PTSD in left and right hemispheres. *Panel A* - ILFlat anatomical connectivity with the anterior part of the superior temporal gyrus in the left hemisphere was significantly different between WTC responders with and without PTSD. *Panel B* - ILFmed connections with the posterior territory of the superior temporal gyrus in the right hemisphere were significantly different among the two groups. *Panel C* - Temporal fibers coursing in the IFOF and representing hippocampal and anterior parahippocampal connections in the right hemisphere were significantly different between WTC responders with and without PTSD. IFOF connections of the posterior part of the right inferior temporal gyrus and the anterior part of the superior temporal gyrus were significantly increased in WTC responders with PTSD. *Panel D* - MdLF connections with the entire superior temporal gyrus in the left hemisphere were significantly increased in WTC responders with PTSD. *Panel E* - Temporal connections in the CB showed a trend towards an increase with hippocampal and parahippocampal cortices but these plastic changes were not significant.

Permutation tests revealed significant group differences across 4 tracts (Table 2), involving the 5 functional hubs identified in Figure 4. These included: lateral ILF fiber, left superior temporal gyrus anterior; MdLF fiber, left superior temporal gyrus anterior and posterior; medial ILF fiber, right superior temporal gyrus; IFOF fiber, right inferior temporal gyrus posterior, parahippocampus anterior, hippocampus and superior temporal gyrus anterior (Table 2). No significant differences were detected in the CB.

**Table 2.** Brain regions in which DAta-driven Network Connectivity Estimate (DANCE) differed between WTC-PTSD and non-PTSD responders.

| Tract | Brain region (ROIs) | Hemisphere | p* |
| --- | --- | --- | --- |
| ILFlat | Superior Temporal Gyrus Ant | LH | 0.036 |
| MdLF | Superior Temporal Gyrus Ant | LH | 0.003 |
|  | Superior Temporal Gyrus Post | LH | 0.022 |
| ILFmed | Superior Temporal Gyrus Post | RH | 0.018 |
| IFOF | Inferior Temporal Gyrus Post | RH | 0.011 |
|  | Parahippocampus Ant | RH | 0.007 |
|  | Hippocampus | RH | 0.005 |
|  | Superior Temporal Gyrus Ant | RH | 0.001 |

White matter tracts and ROIs for which the DATA-driven Network Connectivity Estimate (DANCE) metrics differed significantly between WTC-PTSD and non-PTSD responders. The table reports the white matter tract (Tract), full name of the brain region of interest (ROIs), ROI's hemisphere, and p-value from permutation test. Only p-values obtained from permutation tests and FWE corrected, adjusted for the use of medications (psychotropic and opioids), current depression diagnosis (MDD) and <0.005 are reported.

### WTC Exposure and DAta-driven Network Connectivity Estimate

WTC exposure duration (months on site) moderated the association between PTSD and the DANCE profile in one fiber bundle and one of the nine functional hubs; the IFL medial, right superior temporal gyrus posterior (p-value for interaction = 0.035; Table 3, Figure 5). In WTC-PTSD responders, longer WTC exposure duration was associated with increased DANCE values (95% CI:0.00, 0.05). Results for all remaining tracts, hubs, and models are provided in the Supplementary Materials (Table S1, Figure S3).

**Figure 5.**
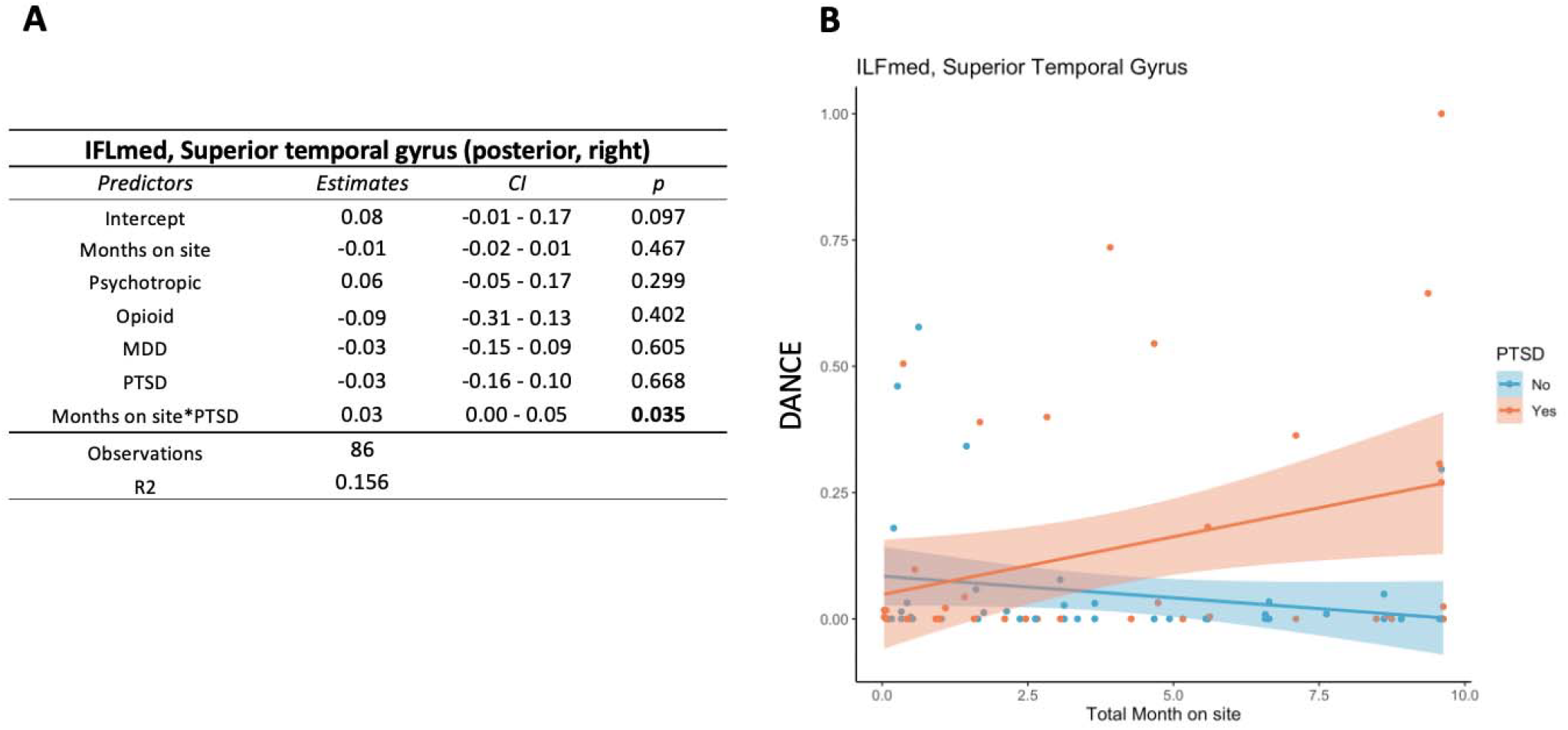
WTC exposure duration moderates DANCE of the AFC tract in the superior temporal gyrus. This table (A) shows WTC exposure duration (months on site) moderates the association between PTSD status and DAta-driven Network Connectivity Estimate (DANCE) profile (p-value for interaction = 0.035, 95% CI [-0.00, 0.05]. (B) This graph plots the relationship (interaction) between WTC exposure duration in months (x-axis) and DANCE profile(y-axis) stratified by WTC-PTSD (orange dots) and non-PTSD (blue dots) for the right posterior parahippocampal gyrus. WTC exposure duration (months on site) moderates the association between PTSD status and DANCE profile in the IFL medial, right superior temporal gyrus posterior (p= 0.035).

## Discussion

We developed and applied a multimodal framework (DANCE) that integrates graph-based functional connectivity metrics with diffusion-based tractography to characterize co-occurring functional-anatomical neuro-profiles in WTC responders with and without PTSD. Several brain regions emerged as key contributors to group differences, serving as functional connectivity hubs, including the bilateral anterior inferior temporal gyrus, right superior parietal lobule, right anterior parahippocampal gyrus, anterior and posterior regions of the right superior temporal gyrus (STG), right caudate nucleus, left amygdala, and brainstem. These findings support a network-level model of PTSD, in which coordinated disruptions in fronto-temporal-limbic circuits underlie alterations in memory processing, emotional regulation, and threat salience.

Our findings build on and extend prior work demonstrating that PTSD is associated with altered brain connectivity^73^. Prior studies have typically reported disruptions in either anatomical or functional networks^74–76^, particularly in temporal and limbic regions involved in memory and emotional processing^77,78^ By jointly modeling functional and structural connectivity, DANCE captures coordinated network-level alterations that are not detectable using single modality approaches.

Consistent with prior literature, these alterations may contribute to heightened stress reactivity and intrusive thoughts or memories^79^. Additionally, hippocampal and parahippocampal regions, essential for memory formation and retention and fear processing, often show impaired connectivity^80^, which can affect how traumatic memories are stored and recalled, leading to the distressing flashbacks characteristic of PTSD. In this context, we identified co-occurring functional-anatomical neuro-profiles that differentiate WTC responders with and without PTSD, achieving a 78% classification accuracy. Using PLS-DA, we observed clear separation in DANCE neuro-profiles between groups and identified key brain hubs that overlapped with those identified previously using permutation testing^11^

These differences were supported by tract-specific alterations across major white matter pathways. DANCE profiles differed between WTC PTSD and non-PTSD in the inferior fronto-occipital fasciculus (IFOF), the medial (ILFmed) and lateral (ILFlat) components of the inferior longitudinal fasciculus, and the middle longitudinal fasciculus (MdLF), implicating connectivity involving the hippocampus, parahippocampus, and temporal cortices. Notably, increased connectivity within ILFmed–STG pathways among PTSD responders suggests altered integration within memory-related circuits, which may reflect maladaptive hyperconnectivity or compensatory reorganization. Together, these findings point to coordinated disruptions in fronto-temporal–limbic networks central to memory, emotion, and threat processing, and highlight candidate circuits for biomarker development and intervention.

Diffusion MRI tractography is the only non-invasive approach for reconstructing major white matter pathways in the human brain. It is most accurate in regions with uniform fiber orientation and less accurate in areas with sparse fibers or complex crossing, especially when used to reconstruct gray matter region-to-region connectivity^81,82^. To mitigate these limitations, we focused our tractography analysis on the core of each white matter tract reducing gyral biases and the confounding influence of superficial fibers. First, we started tracking in the middle of the body of the white matter tract rather than in gray matter. This enables us to avoid artifacts caused by tracking through layers of white matter running parallel to the cortex when initiating the reconstruction of each bundle. Second, we use anatomy-driven masks to minimize the reconstruction of false positives. We then estimated the magnitude or connectional strength between each bundle to each gray matter target. This strategy, validated against primate data, successfully allowed us to overcome the potential challenges posed by in vivo tractography and enabled us to reconstruct both the course and anatomical connections of the white matter tracts linking the functional networks altered in WTC responders who developed PTSD.

We found that DANCE connectional profiles of the four main longitudinal fascicles in the temporal cortex differed significantly between WTC responders with and without PTSD, including both medial and lateral subcomponents of the ILF, while no differences in the cingulum bundle. These findings suggest that PTSD in WTC responders is associated with plastic reorganization of temporal white matter networks, complementing the functional changes previously identified in IFOF, ILF, and MdLF. The convergence of structural and functional abnormalities underscores the clinically relevance of DANCE as a tool for identifying neurobiological signatures of PTSD, though causal links between trauma, functional changes, and long-term anatomical reorganization cannot yet be established. The absence of cingulum differences may indicate that its temporal connections are less vulnerable to trauma, that connectivity normalizes over time, or that PTSD-related changes occur in other brain regions.

The impact of traumatic experiences like WTC exposure on the brain depends on the type of trauma as well as the intensity and duration. In this study, WTC exposure duration exposure duration (months on site) moderated associations between PTSD and DANCE profiles, with longer exposure associated with increased connectivity in the IFLmed-STD among responders with PTSD. These findings suggest that prolonged exposure to traumatic environments may amplify or sustain brain changes, potentially reflecting cumulative neurobiological burden or maladaptive plasticity. This is consistent with prior reports of white-matter alterations and functional reorganization in PTSD following acute, high-intensity trauma and chronic, lower-intensity exposure^83^. Together, findings suggest that the impact of trauma should be understood as a combination of exposure intensity and duration of the traumatic experience, which may contribute to heterogeneity in PTSD phenotypes and help explain variability in affected neural networks across studies. These patterns may also reflect accelerated brain aging processes previously observed in WTC responders^84^.

Limitations of our study include the small sample size and lack of external control group (i.e. non-WTC participants). The sample size precluded splitting the data into independent training and validation sets, raising the possibility of overfitting high-dimensional models. While a strong effort was made to increase representation of women and individuals from underrepresented racial and ethnic groups, further diversity is needed to support subgroup analyses. The age of participants (44-65 years, mean 55) may limit generalizability to younger PTSD populations, although less is known about the effect of PTSD on the brain in individuals in this stage of life and more studies are needed on PTSD and aging. Exposure data were based on retrospective self-report, often collected years after 9/11, and may be subject to recall bias. Additional limitations include the lack of pre-9/11 imaging data, limited information on lifetime trauma history, and lack of a comparison group with subsyndromal, remitted PTSD. While these factors constrain generalizability, prior studies show consistent PTSD-related neural alterations across trauma-exposed populations, supporting the relevance of our findings beyond WTC responders.

The strength of our approach lies in its simultaneous assessment of anatomical and functional connectivity in PTSD. Identifying these multimodal biomarkers is essential for advancing interventions that directly modulate activity in affected neural circuits. Non-invasive techniques such as transcranial magnetic stimulation^85,86^ and transcranial electrical stimulation^87,88^ have shown efficacy in reducing core PTSD symptoms. Emerging approaches, including low-intensity transcranial focused ultrasound stimulation^89,90^ might extend this capability to deeper brain regions, such as the temporal lobe structures identified here, with greater spatial precision. This technique demonstrated the ability to be effective in modulating deep temporal regions in non-human primates and humans^91^. Together, our data-driven connectivity framework provides a unique and promising advancement towards an integrated understanding of the neural mechanisms of PTSD and lays the foundation for more precise, personalized, therapeutic interventions.

## Methods

### Participants

In 2002, the Centers for Disease Control and Prevention (CDC), through the World Trade Center (WTC) Health Program, began monitoring over 50,000 WTC responders using a comprehensive health surveillance protocol at several clinical centers throughout the New York City metropolitan area^26–28^. The Stony Brook University (SBU) site monitors law enforcement and non-law enforcement responders (e.g., construction workers, utility personnel, and volunteers) who primarily reside on Long Island, New York^27,29^. These responders are evaluated for a range of health conditions, including PTSD and major depressive disorder (MDD). A subset of the individuals at the SBU site participated in a voluntary substudy involving structural and functional MRI^27,29–31^. Briefly, all participants were WTC responders involved in rescue, recovery, cleanup, or related operations following the 9/11 attacks^27,28^.

Eligibility criteria included age 44-65 years, English fluency, and meeting MRI safety requirements (e.g., no metal implants, no shrapnel, no claustrophobia, no history of traumatic brain injury, and BMI ≤ 40). During recruitment, case (WTC-PTSD) and control (non-PTSD) groups were matched on age, sex, race/ethnicity, occupation (i.e., law enforcement), and education at the time of 9/11.

### Ethics

Study procedures that follow the Declaration of Helsinki, were approved by the Institutional Review Boards at both Stony Brook University and the Icahn School of Medicine at Mount Sinai. All participants signed informed consents upon explanation of study procedures prior to enrollment.

### WTC exposure duration

All WTC responders completed an exposure assessment questionnaire (EAQ) upon enrollment into the World Trade Center Health Program (WTCHP) General Responder Cohort (GRC). The GRC has been described in detail by Dasaro et al. 2017^27^. All members worked or volunteered on WTC rescue and recovery efforts following the 9/11 attack on the WTC towers, meeting at least one of the following criteria: (i) involved for ≥ 4 hours from September 11 to September 14, 2001, or ≥ 24 hours in September 2001, or ≥ 80 hours from September 11 to July 2002; (ii) member of the Office of the Chief Medical Examiner for New York City, handling and processing human remains for ≥ 4 hours; (iii) worked for the Port Authority Trans Hudson Corporation (PATH), cleaning PATH tunnels for ≥ 24 hours between February 2002 and July 2002.

WTC exposure duration was calculated as time spent (months) of physical presence working on the WTC sites^33,34^. This exposure variable was not available for 10 participants in our study, therefore analyses including this variable included a sample of 86 WTC responders. There is no significant difference between the participants removed (n=10) and the participants included in the analysis (n=86) in any demographic characteristics.

PTSD status (current PTSD or no PTSD) was determined from the Structured Clinical Interview for the DSM-IV (SCID-IV)^32^, administered by trained clinical interviewers, with WTC exposures defined as the index trauma. PTSD symptom subdomains were assessed using SCID-IV subscales reflecting severity of re-experiencing, avoidance, hyperarousal, and negative thoughts (scores ranging from [10-30], [14-42], [10-30], [8-24], respectively). Current MDD (active within the last month) was also assessed using the SCID-IV. MDD, a common comorbidity of PTSD, was not an exclusion criterion.

### MRI and fMRI data acquisition

Magnetic resonance imaging (MRI) and fMRI data acquisition was performed at the Biomedical Engineering and Imaging Institute at Icahn School of Medicine at Mount Sinai using a 3-Tesla SIEMENS mMR Biograph scanner with a 200-channel head and neck coil. For each WTC responder, a high-resolution 3D T1-weighted structural scan was acquired using a MPRAGE sequence (TR = 1900ms, TE = 2.49ms, TI = 900ms, flip angle = 9, acquisition matrix = 256×256 and 224 slices with final voxel size = 0.89×0.89×0.89 mm). Diffusion data were acquired using single shell diffusion-weighted EPO acquisition (TR = 4680ms, TE = 87.6ms, b-value = 1200, 64 diffusion directions, in plane resolution = 2mm, slice thickness = 2mm, matrix size-128*128, multiband factor = 2). Then, functional T2*-weighted, 2D echo planar images were obtained using the following parameters: TR= 1500 ms, TE = 27 ms, pulse angle = 80 degrees, field of view = 22 cm, with a 74 x 74-pixel matrix and a slice thickness of 2.5 mm. Fifty contiguous oblique-axial sections to cover the whole brain where the first four images were discarded to allow the magnetization to reach equilibrium. For each subject, a single 10-minute continuous functional resting state sequence was acquired, for a total of 400 volumes. During resting-state scan, participants were instructed to relax, lay still, not think about anything, with and keep their eyes open. Padding was used for comfort and to reduce head motion.

### Functional MRI data analyses

Image pre-processing, ECM, and statistical analyses were performed using SPM12 (Wellcome Department of Imaging Neuroscience, London, UK), fastECM toolbox^35^ and customized scripts, implemented in MatLab 2016b (The Mathworks Inc., Natick, Massachusetts) and R (v3.4).

### Image preprocessing

Structural MRI data were co-registered and normalized against the Montreal Neurological Institute (MNI) template and segmented into white matter (WM), gray matter (GM) and cerebrospinal fluid (CSF) probability maps. fMRI data were spatially realigned, co-registered to the MNI-152 EPI template, and normalized using the SPM12 segmentation option for EPI images. Normalized data were denoised using ICA-AROMA^36^. Additionally, spatial smoothing was applied (8 millimeters) to the fMRI data. No global signal regression was applied. Using the Harvard-Oxford^37^ atlas, 111 regions of interest (ROI; 48 left and 48 right cortical areas; 7 left and 7 right subcortical regions and 1 brainstem) were defined. For each ROI, time-series were extracted by averaging voxels per time point. Data were “pre-whitened” in a two-step GLM procedure^38,39^ removing the estimated autocorrelation structure. First, raw data were filtered for 6 motion parameters (3 translations and 3 rotations) and resulting residuals were used to estimate and remove autocorrelation using an Auto-Regressive model of order 1 (AR(1)). Realignment parameters, white matter (WM) and cerebrospinal fluid (CSF) signals were removed as confounders from the whitened data.

### Eigenvector Centrality Mapping

Eigenvector centrality mapping (EC) is a graph theory metric that quantifies the influence of a node in a network based on the number of its connections^40^. Fast ECM^35^ was performed on the defined ROI time course data per subject, generating voxel-wise centrality values dependent on summed centrality of neighboring nodes. In the fast ECM toolbox^35^, EC is estimated from the adjacency matrix, which contains the pairwise correlation between the ROIs. To obtain a real-valued EC value, we added +1 to the values in the adjacency matrix. Among possible eigenvectors, only the one associated with the highest eigenvalue (EV) was retained. Subject-level values were averaged at the group level, and high EC values were used to identify influential ROIs, i.e. the hubs^41–43^.

### Diffusion MRI data analyses

Using the direction and magnitude of displacement of water diffusion along the fiber bundles in the brain, diffusion tensor imaging (DTI) estimates metrics reflecting the microstructural properties of whole-brain white matter^44^. Diffusion data were preprocessed with FSL^45^ and the MR Comparative Anatomy Toolbox (MrCat)^46^. Non-diffusion-weighted images were extracted from the full set, averaged, and corrected from spatial RF-field inhomogeneity biases before being linearly registered to the average T1-weighted structural image of the same subject. T1-weighted images were linearly and nonlinearly registered to a study-specific template in FSL MNI-152 space created by averaging images collected from both groups (i.e., WTC-PTSD and non-PTSD). This registration allowed a direct mapping between diffusion-weighted, T1-weighted, and standard space for both WTC-PTSD and non-PTSD responders. Top-up correction was applied to the diffusion-weighted images to correct for susceptibility induced field distortions^45,47^. Current-induced distortions and subject movements were corrected by applying EDDY^48^. Diffusion tensors were fitted to each voxel using DTIFIT^49^. Voxel-wise crossing fiber model fitting of diffusion orientations was performed for all subjects (control and WTC responders) using FSL’s BedpostX^50^. A multi-shell extension was used to reduce overfitting of crossing fibers due to non-monoexponential diffusion decay^51,52^. Up to three fiber orientations per voxel were allowed. This produced voxel-wise posterior distributions of fiber orientations that were subsequently used in probabilistic tractography.

### Probabilistic tractography

Based on diffusion MRI data, probabilistic tractography reconstructs the microstructural features of white matter tracts and maps the brain’s structural connection at macroscale level^53^. Building on our previous research identifying functional hubs within the temporal cortex in WTC-PTSD compared to non-PTSD responders,^11^ we examined five major white matter tracts interconnecting these regions the temporal branch of the cingulum bundle (CB), the medial (IFLmed) and lateral (IFLlat) projections of the inferior longitudinal fascicle, the middle longitudinal fascicle (MdLF) and the inferior-fronto-occipital fascicle (IFOF).

Tractography recipes for all tracts were defined with references to published atlases^54,55^, tract-tracing studies^56–58^, and diffusion tractography studies^24,59–62^. Masks were created in MNI space and warped to individuals’ diffusion space. The CB seed mask was drawn in the white matter medial and ventral to the hippocampus to capture temporal connections. The ILF was segmented into medial and lateral components following Latini et al., (2017)^63^ and Roumazeilles et al., (2020)^62^ with the ILFmed seeded in the white matter of the medial occipitotemporal and the ILFlat seeded laterally within the lateral occipitotemporal. The MdLF was seeded with the white matter occupying the middle temporal gyrus, comprising the inferior temporal and superior temporal sulcus. The IFOF seed mask was placed in the superior temporal gyrus white matter. Seed, exclusion, and waypoint masks were created on the template image and warped back to individuals for probabilistic tractography analysis in subject-space.

Probabilistic tractography was performed in individuals diffusion space with identical seeding and tracking parameters across tracts and groups (WTC-PTSD; non-PTSD). Parameters included: maximum of 3200 steps per sample; 10,000 samples; step size of 0.1 mm; and curvature threshold of 0.2.

A visualization map or ‘tractogram’ was constructed for each participant, to allow comparison across tracts, subjects, and groups^24,64^. Each tractogram was log-transformed to account for the exponential decrease of visitation probability with distance and normalized by dividing each voxel’s value by the 75th percentile value across the tractogram, reducing biases due to differences in the number of streamlines across tracts. Analyses focused on comparing temporal white matter tracts between WTC-PTSD and non-PTSD responders. Subject-specific temporal streamlines were warped back to template space and averaged into tract-specific group tractograms, with group centers defined as the mean of individual tractograms. For visualization only, normalized tractograms were thresholded with minimum and maximum values equal to 0.5 and 2, respectively.

### Data-driven Network Connectivity Estimate: A Multimodal MRI approach

The DAta-driven Network Connectivity Estimate (DANCE) is a multimodal connectivity metric that quantifies the correspondence between white matter streamline based on structural connectivity and functional network hubs^23^. DANCE integrates complementary MRI modalities to characterize how brain structure and function interact. Specifically, it combines (1) resting-state functional MRI (rs-FMRI) to identify brain regions exhibiting altered neural activity (i.e., functional hubs), and (2) diffusion-weighted MRI tractography to map the underlying anatomical connections between these hubs. This multimodal framework provides a data-driven measure of the coupling between functional organization and structural connectivity across the brain^24^. We focused on connected (or disconnected) functional hubs that influence global information flow and integration. Using probabilistic tractography, we reconstructed the temporal fiber bundles connecting the functional hubs. This allowed us to test the hypothesis that WTC-PTSD responders exhibit concurrent changes in functional and anatomical connectivity within the same brain networks. We generated anatomical connectivity fingerprints for each tract and quantitatively compared them between WTC-PTSD and non-PTSD responders^65–67^. Therefore, DANCE identifies co-occurring functional and anatomical connectivity alterations in key brain networks affected by PTSD, offering insight into the neural circuitry underlying the disorder and potential biomarkers for future research and intervention.

Once functional hubs were estimated from resting-state fMRI, we quantified the distribution of streamlines connecting these hubs by reconstructing the tract-specific connectivity fingerprint^23,68^. Such fingerprints can be used to compare tracts’ temporal projections with one another and to compare the tracts across groups (i.e., WTC-PTSD versus non-PTSD responders). Temporal ROIs were defined using published coordinates (Harvard-Oxford Atlas in FSL), created on the group average template in MNI space, and warped back to subject-specific space for tractography. The white matter/gray matter border of each ROI was then extracted in each subject and used as the parcel in the anatomical connectivity fingerprint. This approach allows us to maximize the estimation of streamlines projecting to each region while minimizing the reduction of signal consequent to the tissue-related poor anisotropic signal characteristic of ROIs created in gray matter only. For each tract, fingerprints are constructed by averaging the number of streamlines hitting each ROI. To allow comparison across the two groups, these values were normalized by dividing each tract-fingerprint by its maximum values across the selected ROIs. The center of the group is defined as the mean of the individual subject connectivity fingerprints. We compared connectivity fingerprints by calculating the Manhattan distance between them^46^. The overall pipeline is demonstrated in Figure 1.

## Statistical analyses

### Predictive modeling

To model the divergence of EC in WTC-PTSD and non-PTSD responders’ values in 111 ROIs, we applied partial least squares discriminant analysis (PLS-DA)^69,70^. Using EC values as inputs, this supervised dimensionality-reduction technique constructs a lower-dimensional subspace that maximizes the separation between WTC-PTSD and non-PTSD responders, emphasizing predictive efficacy rather than statistical significance. A receiver operating characteristic (ROC) curve was generated to predict WTC-PTSD from the first discriminant axis with varying classification thresholds^71^. To quantify uncertainty in the ROC curve and discriminant prediction, a confidence interval of the ROC curve was computed using 2000 stratified bootstraps. We implemented a rank-based statistical test to determine if the performance of our classification algorithm was significantly different than a classification made randomly. P-values derived from this method are interpreted as the model’s overall predictive performance. The loadings of each region on a given dimensional subscale were evaluated to determine which regions contributed most to the overall divergence between WTC-PTSD and non-PTSD responders (i.e., functional hubs).

Influential functional hubs were defined as ROIs with the highest discriminant loadings and those differing significantly between groups using a standard statistical approach (Invernizzi et al, 2022) ^11^. PLS-DA models were implemented in R using the mixOmics package^72^.

We quantify DANCE profile differences between groups by permuting labels with 10000-time repetitions. ROIs per anatomical tract with p≤ 0.05 were considered statistically significant. Family wise error (FWE) correction was applied for the number of group level comparisons and for the total number of ROIs and tracts analyzed. Only FWE corrected p-values are reported.

### Hypothesis Testing

To test whether DANCE profiles moderated the association between WTC exposure duration and WTC-PTSD, generalized linear models (GLM) were applied using current PTSD diagnosis and cumulative WTC exposure (months) as predictors and DANCE profiles for each region as outcomes. Models were adjusted for current psychotropic/opioids use and current depression. Only EC values of hubs in permutation analyses were included as outcomes. GLMs were implemented using R (Version 1.4.1717).

## Data Availability

All data produced in the present study are available upon reasonable request to the authors

## Acknowledgements

The authors would like to acknowledge support from the Centers for Disease Control and Prevention for supporting the neuroimaging study (CDC/NIOSH U01 OH011314), the National Institute on Aging that supports research on characterization and treatment of Alzheimer’s disease (NIH/NIA P50 AG005138), and aging-related work in this population (NIH/NIA R01 AG049953). We would also like to acknowledge ongoing funding to monitor World Trade Center responders as part of the WTC Health and Wellness Program (CDC 200-2011-39361).

## Supplementary material

**Figure S1.**
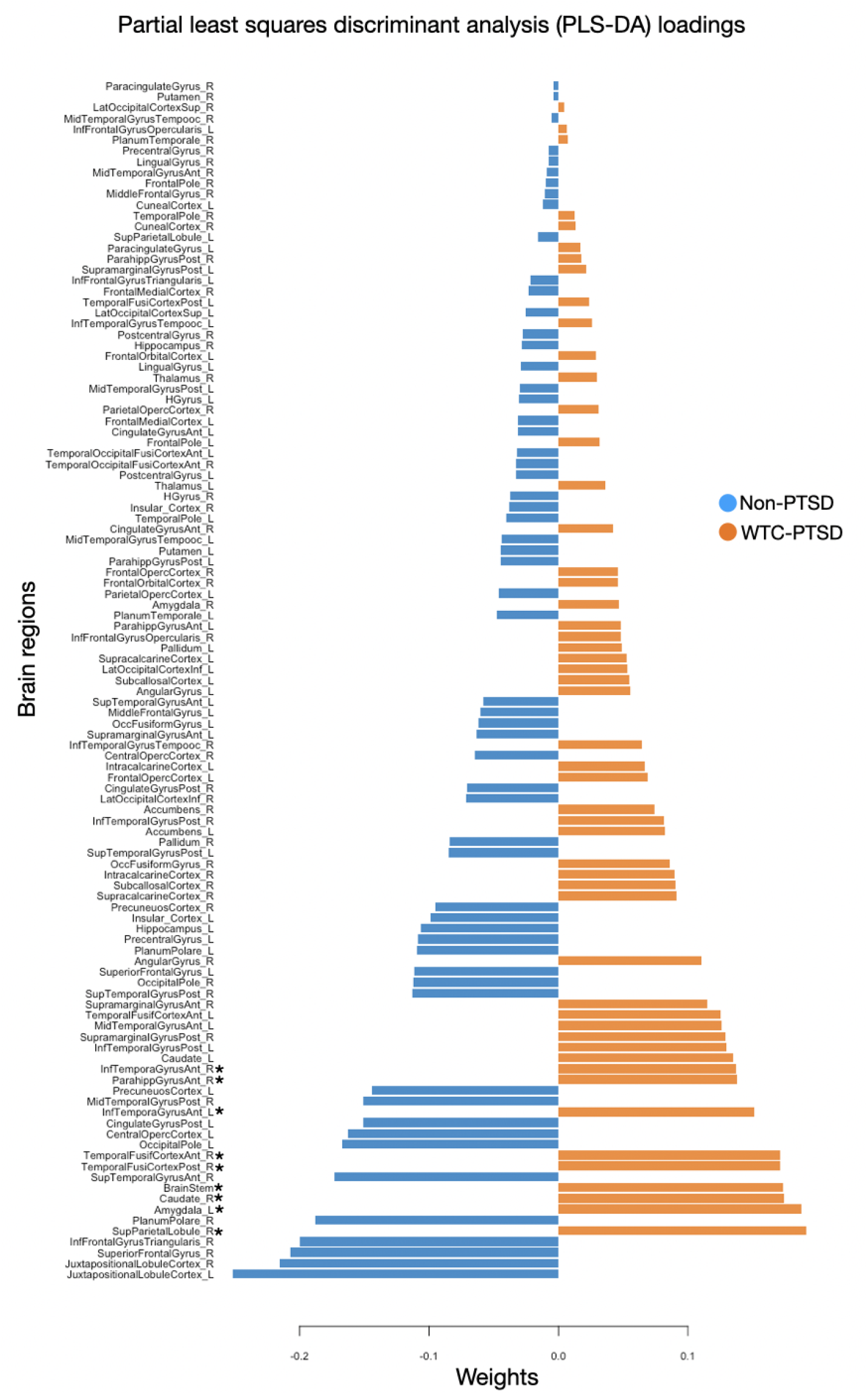
Partial least squares discriminant analysis (PLS-DA) loadings. The partial least squares discriminant analysis (PLS-DA) identifies the loading of each brain region (n=111) on a given dimensional subscales to determine which regions contribute most to the overall divergence between WTC responders with and without PTSD. Loadings are color coded to indicate the group with a higher mean score on that variable, blue lines indicating measures where WTC-responders with no PTSD, and orange lines indicating variables where WTC responders with PTSD. The loadings direction (positive or negative) indicate the direction of the associations between single brain regions and PTSD. * highlights brain areas that result also significantly different across groups using the standard statistical approach (see Methods and Result sections for more details).

**Figure S2.**
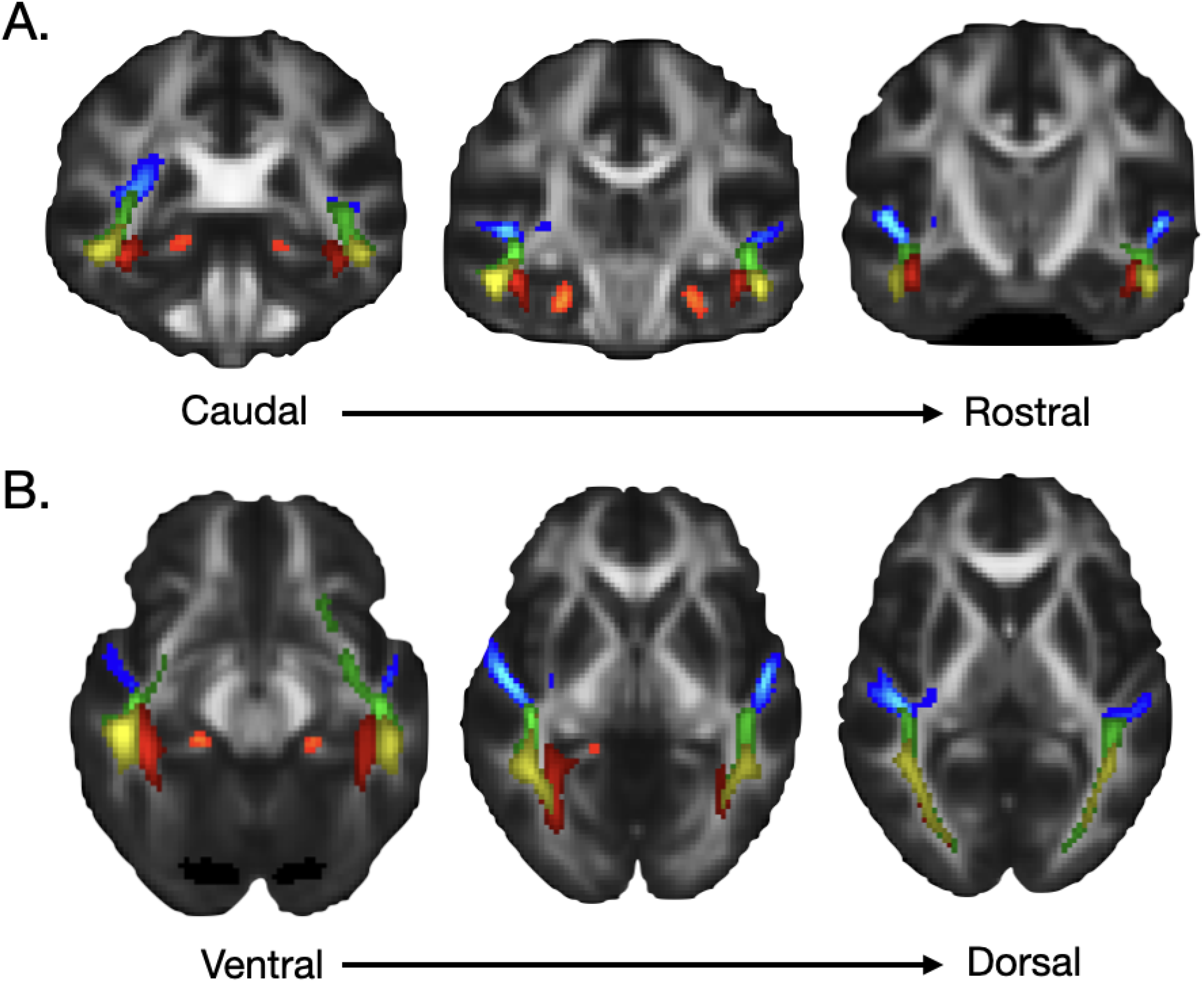
Tractography analysis. Five white matter tracts were defined using DTI data: the temporal branch of the cingulum bundle (CB), the medial (ILFmed) and lateral (ILFlat) projections of the inferior longitudinal fascicle, the middle longitudinal fascicle (MdLF) and the inferior-fronto-occipital fascicle (IFOF).

**Figure S3.**
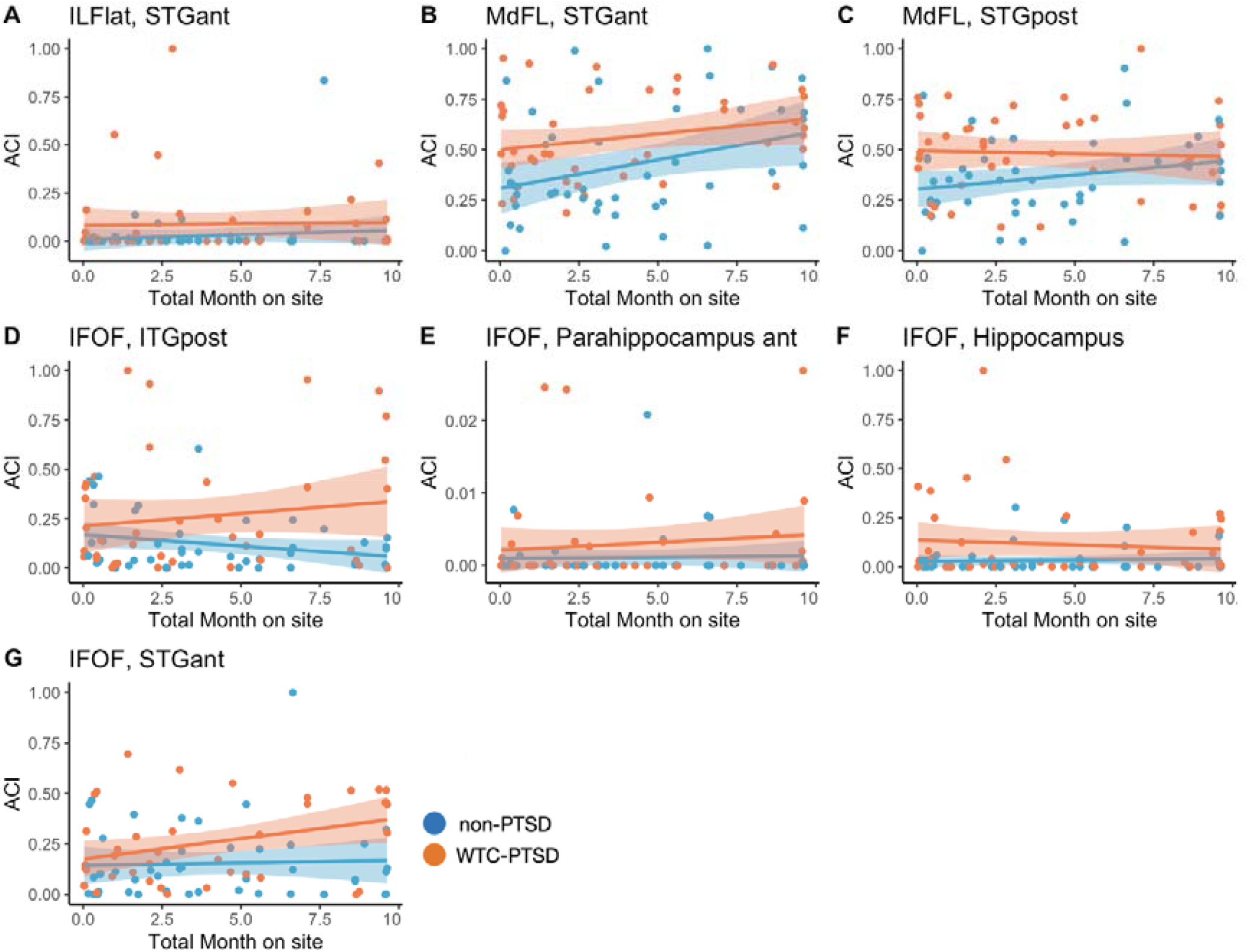
Effects of WTC exposure duration on centrality values for the identified hubs. These panels show the relation between WTC exposure duration expressed in months and eigenvector centrality values for the nine identified hubs. Orange and blue dots represent WTC-responders with and without PTSD respectively.

**Table S1.** Association between EC values, PTSD status and WTC exposure duration. Generalized regression models (GLM) examining WTC exposure duration (i.e., months on site) moderates the association between PTSD (WTC-PTSD vs non-PTSD) and EC values controlling for major depressive disorder (MDD) and medication use (psychotropic and opioid) on eigenvector (EC) value of a single brain area (defined using the Harvard-Oxford atlas).

| Predictors | ILFlat, STGant LH |  |  | MdFL, STGant LH |  |  | MdFL, STGpost LH |  |  | IFOF, ITGpost RH |  |  | IFOF, Parahipp Ant RH |  |  | IFOF, Hippocampus RH |  |  | IFOF, STGant RH |  |  |
| --- | --- | --- | --- | --- | --- | --- | --- | --- | --- | --- | --- | --- | --- | --- | --- | --- | --- | --- | --- | --- | --- |
|  | Estimates | CI | p | Estimates | CI | p | Estimates | CI | p | Estimates | CI | p | Estimates | CI | p | Estimates | CI | p | Estimates | CI | p |
| (Intercept) | 0.002 | -0.076 – 0.081 | 0.954 | 0.313 | 0.196 – 0.431 | **<0.001** | 0.305 | 0.204 – 0.405 | **<0.001** | 0.167 | 0.054 – 0.280 | **0.004** | 0.001 | -0.002 – 0.004 | 0.467 | 0.025 | -0.048 – 0.098 | 0.500 | 0.137 | 0.044 – 0.230 | **0.004** |
| TOTALMONTHONSITE | 0.006 | -0.009 – 0.022 | 0.413 | 0.029 | 0.007 – 0.052 | **0.011** | 0.015 | -0.004 – 0.035 | 0.129 | -0.010 | -0.032 – 0.012 | 0.354 | 0.000 | -0.000 – 0.001 | 0.876 | 0.002 | -0.012 – 0.016 | 0.763 | 0.005 | -0.013 – 0.023 | 0.603 |
| npsych | 0.049 | -0.047 – 0.145 | 0.313 | -0.028 | -0.171 – 0.115 | 0.705 | 0.005 | -0.118 – 0.128 | 0.934 | 0.008 | -0.130 – 0.146 | 0.909 | -0.000 | -0.003 – 0.003 | 0.981 | 0.013 | -0.076 – 0.102 | 0.780 | 0.045 | -0.068 – 0.158 | 0.436 |
| opioid | -0.057 | -0.247 – 0.134 | 0.561 | -0.167 | -0.451 – 0.117 | 0.250 | -0.073 | -0.317 – 0.171 | 0.556 | -0.070 | -0.345 – 0.205 | 0.617 | -0.001 | -0.007 – 0.006 | 0.791 | -0.055 | -0.232 – 0.122 | 0.543 | -0.125 | -0.351 – 0.100 | 0.275 |
| current MDD | 0.076 | -0.027 – 0.179 | 0.150 | 0.099 | -0.055 – 0.253 | 0.207 | -0.023 | -0.154 – 0.109 | 0.737 | -0.017 | -0.165 – 0.132 | 0.827 | -0.000 | -0.004 – 0.003 | 0.825 | 0.021 | -0.075 – 0.117 | 0.670 | 0.080 | -0.042 – 0.201 | 0.200 |
| PTSD [Yes] | 0.045 | -0.069 – 0.159 | 0.443 | 0.160 | -0.010 – 0.330 | 0.064 | 0.198 | 0.052 – 0.344 | **0.008** | 0.052 | -0.112 – 0.216 | 0.537 | 0.001 | -0.003 – 0.005 | 0.505 | 0.101 | -0.005 – 0.207 | 0.061 | 0.002 | -0.133 – 0.137 | 0.975 |
| TOTALMONTHONSITE × PTSD [Yes] | -0.008 | -0.030 – 0.014 | 0.450 | -0.013 | -0.046 – 0.019 | 0.419 | -0.018 | -0.046 – 0.010 | 0.212 | 0.023 | -0.009 – 0.055 | 0.155 | 0.000 | -0.001 – 0.001 | 0.624 | -0.008 | -0.028 – 0.013 | 0.461 | 0.012 | -0.014 – 0.038 | 0.358 |
| Observations | 86 |  |  | 86 |  |  | 86 |  |  | 86 |  |  | 86 |  |  | 86 |  |  | 86 |  |  |
| R<sup>2</sup> | 0.086 |  |  | 0.187 |  |  | 0.110 |  |  | 0.116 |  |  | 0.041 |  |  | 0.092 |  |  | 0.160 |  |  |

## Notes

<u>Conflict of interest:</u> None.

### Competing Interest Statement

The authors have declared no competing interest.

